# Genetically Proxied NLRP3 Inflammasome Downregulation and Cardiovascular Disease

**DOI:** 10.64898/2026.11.16.26368176

**Authors:** Benjamin P Zuckerman, Panagiotis Zangas, Lanyue Zhang, Anastasiia Osadcha, Sizheng Steven Zhao, Marios K Georgakis

## Abstract

**Background:** The inhibition of NOD-, LRR- and pyrin domain-containing protein 3 (NLRP3) inflammasome is an emerging therapeutic strategy for atherosclerotic cardiovascular disease (ASCVD). Although early clinical trials demonstrate reductions in inflammatory biomarkers, whether these translate into cardiovascular benefit remains uncertain. We investigated the potential efficacy and safety of NLRP3 inhibition using human genetics.

**Methods:** We developed a *cis*-genetic instrument for NLRP3 downregulation using C-reactive protein (CRP) associated variants (P<5×10⁻⁸) within the *NLRP3* locus (±100 kb of *NLRP3* transcription start site) from 575,531 participants. Mendelian Randomization and colocalization analyses assessed the effect of the genetic instrument against gene expression of *NLRP3* and other inflammatory mediators in immune cells, downstream inflammatory biomarkers, and pharmacodynamic responses to the NLRP3 inhibitor ruvonoflast. Two-sample Mendelian randomization assessed cardiovascular, cardiometabolic, and infectious outcomes, followed by phenome-wide analyses of 1,387 clinical outcomes.

**Results:** Using 14 variants in the *NLRP3* locus, we constructed a genetic instrument that corresponded to a 19% difference in circulating CRP between the extreme percentiles of its distribution. Colocalisation evidence suggested the effects on CRP to be driven by reduced *NLRP3* expression in naïve monocytes (PP.H4=98.7%). In accordance with the expected biological role of NLRP3, genetically proxied NLRP3 downregulation by the constructed instrument was associated with lower IL-18 (β=−0.11; 95%CI, −0.14 to −0.08) and IL-6 (β=−0.15; 95%CI, −0.17 to −0.12) and broadly similar pharmacodynamic effects to the NLRP3 inhibitor ruvonoflast across 8 biomarkers. Genetically proxied NLRP3 inhibition was associated with lower risk of recurrent pericarditis (OR per 19% decrease in CRP, 0.68, 95%CI, 0.58-0.80), but slightly elevated risk of coronary artery disease (OR, 1.06; 95%CI, 1.03–1.08) and peripheral arterial disease (OR, 1.10; 95%CI, 1.04–1.15) and no effect on ischaemic stroke (OR, 0.98; 95%CI, 0.94-1.02). The instrument was also associated with higher carotid-intima-media thickness and a worse cardiometabolic profile with increasing effects on blood pressure, waist-to-hip ratio, LDL cholesterol, and triglycerides. We found no associations with increased risk for prespecified infectious disease outcomes.

**Conclusions:** A genetic proxy for NLRP3 inflammasome downregulation reproduced key molecular effects of pharmacological NLRP3 inhibition but was not associated with reduced ASCVD risk.

**Clinical perspective:** *What is new?:* - We developed a genetic proxy of NLRP3 inflammasome downregulation comprising 14 variants within the *NLRP3* locus. - Mirroring the biology of NLRP3 signaling, genetically proxied NLRP3 downregulation was associated with lower IL-6 and IL-18 concentrations and showed broadly concordant biomarker effects with the oral NLRP3 inhibitor, ruvonoflast. - Despite mimicking pharmacologic NLRP3 inhibition at a biomarker level, genetically proxied NLRP3 downregulation was not associated with reduced lifetime cardiometabolic risk.

*What are the clinical implications?:* - In view of emerging therapeutic efforts to target NLRP3 inflammasome, we could not find human genetic evidence supporting that NLRP3 downregulation could reduce risk of cardiovascular events.

## Introduction

Cardiovascular disease (CVD) remains the leading cause of death worldwide^1^. The projected increase in its burden over the coming decades highlights the need for novel preventive strategies^2,3^. Targeting vascular inflammation has emerged as one therapeutic avenue for CVD prevention^4^. Among inflammatory pathways, the interleukin (IL)-1β–IL-6 axis received particular attention, supported by converging experimental, genetic, and clinical evidence. Proof of concept for this pathway was established in the CANTOS trial, in which inhibition of IL-1β with canakinumab reduced major cardiovascular events^5–7^.

Canakinumab was not developed further for atherosclerotic cardiovascular disease (ASCVD), but attention shifted toward upstream and downstream molecules. Downstream of IL-1β, three monoclonal antibodies targeting IL-6^8–10^ successfully completed phase 2 trials. However, ziltivekimab, one of these agents, failed to demonstrate cardiovascular benefit in the phase 3 ZEUS trial in patients with ASCVD and chronic kidney disease^11^.

Upstream of both IL-1β and IL-6 lies the NOD-, LRR- and pyrin domain-containing protein 3 (NLRP3) inflammasome, a central intracellular sensor of cellular stress that is activated by diverse danger signals, including cholesterol crystals within atherosclerotic plaques^12^.

NLRP3 activation triggers the maturation and release of several pro-inflammatory cytokines, most notably IL-1β and IL-18. By acting upstream of the IL-1β–IL-6 axis, NLRP3 inhibition offers the potential to suppress multiple downstream inflammatory pathways simultaneously. Although clinical development is less advanced than for IL-6 inhibitors, several NLRP3 inhibitors are progressing rapidly. Recently, the oral small-molecule inhibitor ruvonoflast reduced high-sensitivity C-reactive protein (hsCRP), IL-6, and fibrinogen concentrations in patients with high hsCRP levels at elevated cardiovascular risk^13^. Similar reductions in hsCRP have also been reported for additional oral NLRP3 inhibitors (BGE-102, parunoflast and MRT-8102) in early-stage trials of patients with an unfavorable cardiometabolic profile^14–16^. Whether these biomarker improvements translate into reductions in cardiovascular events, however, remains unknown.

Human genetic studies played a central role in prioritising IL-6 signaling as a therapeutic target. Naturally occurring variants in the genes encoding IL-6^17^ or its receptor^18–26^ that attenuate downstream signaling are consistently associated with lower lifetime risk of CVD, providing genetic support for pharmacological IL-6 inhibition. By contrast, there have been only limited efforts to identify genetic proxies of NLRP3 inhibition that could be used to evaluate this emerging therapeutic strategy. One previous study reported that a common intronic NLRP3 variant increased *NLRP3* expression and cardiovascular mortality^27^, but these findings were not replicated in subsequent genome-wide association studies (GWAS) of coronary artery disease^28^ or other cardiovascular phenotypes^29,30^.

Importantly, inhibition of NLRP3 may not simply recapitulate the effects of inhibiting downstream cytokines such as IL-1β or IL-6. Experimental studies suggest that *Nlrp3* haploinsufficiency in mice induces compensatory overexpression of *Nlrp1*, potentially accelerating inflammation and aging over time^31^. In addition, targeting different levels of the inflammatory cascade may have distinct safety profiles. For example, IL-1β inhibition with canakinumab was associated with an increased risk of fatal infections in CANTOS^5–7^, highlighting the need to better understand the efficacy and safety implications of targeting NLRP3 itself.

To address these gaps, we developed a genetic instrument proxying NLRP3 inflammasome downregulation by leveraging variants in the locus of the gene encoding NLRP3 and used it to predict the effects of pharmacological NLRP3 inhibition on cardiometabolic and safety outcomes. We first validated the instrument against the expected biological effects of NLRP3 inhibition and biomarker changes observed in the recently completed phase 1b trial of the oral NLRP3 inhibitor ruvonoflast. We then evaluated its associations with cardiovascular and metabolic diseases, as well as infectious disease outcomes.

## Methods

### Study Design

We performed a drug-target Mendelian randomization (MR) study^32^ to evaluate the effects of genetically proxied NLRP3 inflammasome perturbation on downstream inflammatory biomarkers, cardiometabolic outcomes, and infectious diseases. Genetic instruments comprised C-reactive protein (CRP)-associated cis-acting variants within the *NLRP3* locus. Circulating CRP was selected as the biomarker for instrument construction because it represents a distal, clinically validated marker of NLRP3–IL-1β–IL-6 pathway activity and is an established pharmacodynamic readout in clinical trials of NLRP3 inhibition^13^. The study design is summarized in **Figure 1**.

**Figure 1.**
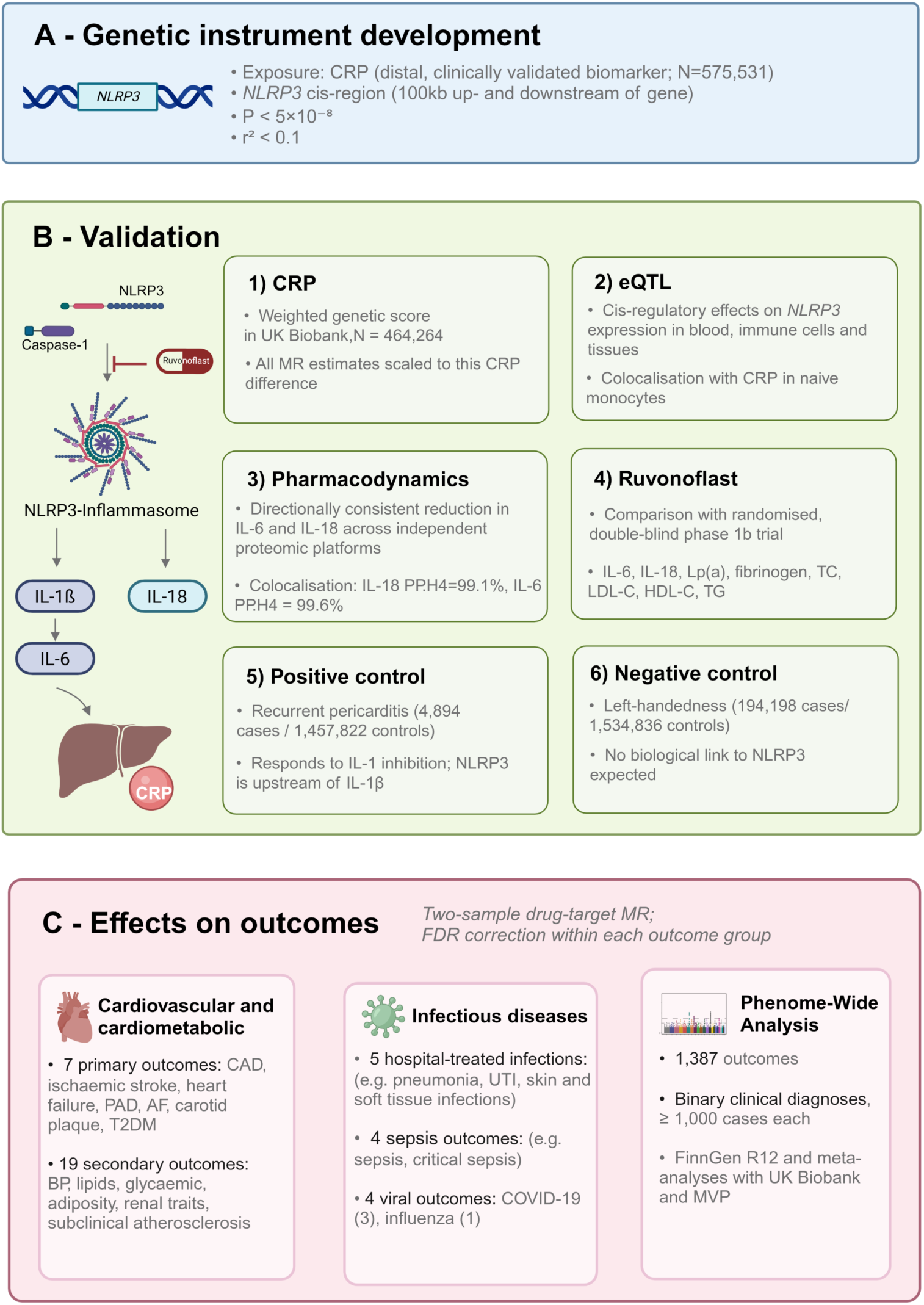
Overview of the study design. **(A)** Construction of the genetic instrument proxying NLRP3 inflammasome downregulation. **(B)** Instrument validation analysis description (genetic score calculation, colocalization with naïve monocyte eQTL, pharmacodynamic validation and comparison with clinical trial results, positive and negative control analyses), **(C)** Drug-target two-sample MR analysis with cardiovascular diseases, cardiometabolic traits and infections as outcomes, followed by a phenome-wide association study. CRP: C-reactive protein, NLRP3: NOD-, LRR- and pyrin domain-containing protein 3, MR: Mendelian Randomization, IL-6: interleukin-6, IL-18: interleukin-18, Lp(a): lipoprotein(a), TC: total cholesterol, LDL-C: low-density lipoprotein cholesterol, HDL-C: high-density lipoprotein cholesterol, TG: triglycerides, IL-1ß: interleukin-1 beta, CAD: coronary artery disease, PAD: peripheral arterial disease, AF: atrial fibrillation, T2DM: type 2 diabetes mellitus, BP: blood pressure, UTI: urinary tract infection, MVP: Million Veteran Program. Created in https://www.biorender.com

### Data Sources

Genome-wide association study (GWAS) summary statistics for circulating CRP were obtained from Said et al^33^, comprising 575,531 predominantly European-ancestry participants. Summary statistics for inflammatory biomarkers, cardiovascular and cardiometabolic outcomes, infectious diseases, quantitative cardiovascular traits, and circulating proteins were obtained from publicly available GWASs conducted predominantly in individuals of European ancestry. Full details of the contributing GWASs, including sample size, ancestry and accession identifiers, are provided in **Supplementary Table S1.**

### Construction of the NLRP3 Genetic Instrument

Genetic proxies for NLRP3 perturbation were identified as CRP-associated variants within the *NLRP3* cis-region (GRCh37; chromosome 1:247,579,458–247,612,410). Variants within ±100 kb of *NLRP3* were selected at genome-wide significance (P<5×10⁻⁸) and clumped for linkage disequilibrium (LD) at r²<0.1 using the European subset of the 1000 Genomes Project Phase 3 reference panel^34^ to construct the primary instrument. Robustness to the choice of cis-window was assessed using a ±10-kb window. A more stringent LD-clumping threshold (r² < 0.01) was additionally applied to assess the robustness of estimates to residual linkage disequilibrium among genetic instruments. The proportion of variance in circulating CRP explained by each selected genetic variant (R²) was calculated^35,36^, and the instrument strength was evaluated using F statistics^35–37^.

### Genetic Score Construction and Effect Scaling

To express the effect of genetically proxied NLRP3 inhibition on a clinically interpretable scale, MR estimates were rescaled to the difference in circulating CRP observed between the extremes of the distribution of a weighted genetic score comprising the selected genetic variants. This was performed in the UK Biobank (N = 464,264) and methodology was described elsewhere.^17,38^. Scaling was applied only to the final MR estimates. Scaled estimates were subsequently re-oriented to represent the effect of genetically proxied NLRP3 downregulation.

### Functional Validation

Instrument variants were additionally annotated using the Ensembl Variant Effect Predictor (VEP)^39^ against the *NLRP3* MANE Select transcript to characterise their predicted functional consequences. The functional validity of the *NLRP3* instrument was evaluated using complementary expression quantitative trait locus (eQTL) resources, including the eQTL Catalogue^40^ and eQTLGen^41^. Nominally significant (two-sided *P*<0.05) eQTL associations were used to descriptively characterise the tissue- and cell-specific regulatory landscape of the *NLRP3* locus. Given the predominant role of the NLRP3 inflammasome in myeloid cells, particularly monocytes and macrophages^42,43^, colocalization analyses were then performed to assess whether *NLRP3* eQTL signals in relevant myeloid lineages, including naïve monocytes^44^, and circulating CRP shared a causal variant.

### Pharmacodynamic Validation

To evaluate whether genetically proxied NLRP3 perturbation reproduced the expected downstream inflammatory effects of NLRP3 inhibition, we examined associations of the primary instrument with circulating IL-6 (N=76,592) and IL-18 (N=76,646), using the largest independent proteomic platform per biomarker as the primary analysis, across multiple independent proteomic platforms^45–50^. IL-18 is directly matured following NLRP3-mediated caspase-1 activation, whereas IL-6 represents a downstream mediator of IL-1β signaling, thereby capturing complementary components of the NLRP3 inflammatory pathway^43^.

Although IL-1β is itself a canonical downstream product of NLRP3 inflammasome activation, its low circulating abundance due to short half-life^51^ presents challenges for reliable quantification in large-scale proteomic datasets. As such, it was not available in the primary proteomic dataset^45^. To further assess concordance with pharmacological NLRP3 inhibition, MR-derived biomarker effects were compared with pharmacodynamic responses reported in a randomized phase 1b trial of the NLRP3 inhibitor ruvonoflast^13^. Comparisons included IL-6, IL-18, fibrinogen, lipoprotein(a), total cholesterol, LDL cholesterol, HDL cholesterol, and triglycerides^45–49,52^ **(Supplementary Table S1)**. Genetic estimates, expressed in outcome-specific standard deviation (SD) units, were selected and compared with the corresponding pharmacological effects of ruvonoflast. The effects of ruvonoflast were calculated as placebo-adjusted percentage changes in mean biomarker levels from baseline to day 28. Concordance between genetic and pharmacological effects were assessed using Spearman rank correlation.

### Positive and Negative Control Analyses

Recurrent pericarditis (4,894 cases/1,457,822 controls)^53^ was selected as a positive-control outcome given the established efficacy of IL-1 inhibition in this disease^54,55^ and the role of NLRP3 upstream of IL-1β activation^56^. Left-handedness (194,198 cases/1,534,836 controls)^57^ was selected as a negative-control outcome.

### Cardiometabolic Analyses

Cardiovascular and cardiometabolic outcomes were prespecified to evaluate the potential clinical effects of genetically proxied NLRP3 inhibition. Primary outcomes comprised coronary artery disease (CAD; 181,522 cases/984,168 controls)^28^, ischaemic stroke (62,100 cases/1,234,808 controls)^30^, heart failure (139,533 cases/1,568,809 controls)^29^, peripheral arterial disease (PAD; 31,307 cases/211,753 controls)^58^, atrial fibrillation (228,926 cases/1,611,415 controls)^59^, carotid plaque (21,540 cases/26,894 controls)^60^, and type 2 diabetes mellitus (428,452 cases/2,107,149 controls)^61^. Secondary exploratory outcomes were selected to characterise effects on intermediate cardiovascular and cardiometabolic phenotypes and comprised measures of subclinical atherosclerosis (coronary artery calcification^62^, carotid intima-media thickness^60^), haemodynamic traits (systolic and diastolic blood pressure and pulse pressure^63^); adiposity (body mass index and waist-to-hip ratio adjusted for BMI^64^); glycaemic traits (HbA1c, fasting glucose, and fasting insulin^65^); lipid traits (HDL cholesterol, LDL cholesterol, non-HDL cholesterol, total cholesterol, and triglycerides^52^); and renal traits (blood urea nitrogen [BUN], estimated glomerular filtration rate [eGFR], cystatin C-based eGFR^66^ and urine albumin-to-creatinine ratio^67^) **(Supplementary Table S1)**.

### Infectious Disease Safety Analyses

We evaluated 13 prespecified infectious disease outcomes. Hospital-treated infections comprised any infection (123,508 cases/577,141 controls), pneumonia (55,241/645,408), skin and soft tissue infection (29,247/671,402), urinary tract infection (21,958/464,526) and Candida infection (7,286/690,977). Sepsis outcomes comprised sepsis requiring hospital admission (18,931/663,531), sepsis in individuals aged <75 years (11,568/462,869), critical sepsis (1,380/429,985) and 28-day mortality following critical sepsis (347/431,081). Viral infection outcomes comprised COVID-19 (122,616/2,475,240), COVID-19 requiring hospitalisation (32,519/2,062,805), and very severe respiratory COVID-19 (13,769/1,072,442), each compared with the population, and influenza (5,133/429,900). Data were drawn from UK Biobank and FinnGen^68,69^, the COVID-19 Host Genetics Initiative^70^, and the Million Veteran Program^71^ **(Supplementary Table S1)**. Where the same phenotype was available from multiple sources, the GWAS with the largest sample size was selected for the primary analysis.

### Statistical analysis

Two-sample MR analyses were performed using harmonised exposure and outcome summary statistics. Fixed-effects inverse-variance weighted (IVW) MR was prespecified as the primary estimator^72^. Robustness was assessed using multiplicative random-effects IVW, weighted median^73^, MR-Egger regression^74^, MR-PRESSO^75^, and leave-one-out analyses. The Cochran’s Q test was used to assess between-variant heterogeneity^76^.

Following MR estimation, effect estimates were re-oriented to represent genetically proxied NLRP3 downregulation and rescaled to the corresponding genetically predicted reduction in circulating CRP, as described above.

Regional colocalisation analyses were performed using the approximate Bayes factor implementation of coloc, with prior probabilities of association with trait 1 (p₁=1×10⁻⁴), trait 2 (p₂=1×10⁻⁴), and both traits through a shared causal variant (p₁₂=1×10⁻⁵)^77^. Analyses were conducted within a ±100-kb window around the *NLRP3* locus. Colocalisation was assessed using the posterior probability of a shared causal variant (PP.H4) and the conditional posterior probability of a shared causal variant given association with both traits (PP.H4/[PP.H3 + PP.H4]). For each measure, a posterior probability >0.80 was considered strong evidence of colocalisation, whereas >0.50 was considered suggestive evidence. Posterior probabilities for all hypotheses (PP.H0–PP.H4) are reported in the **Supplement.**

Benjamini–Hochberg false discovery rate (FDR) correction was applied separately within the prespecified biological validation outcomes, binary cardiometabolic outcomes, continuous cardiometabolic traits, infectious disease safety outcomes, and phenome-wide outcomes^78^. MR sensitivity analyses were considered assessments of robustness rather than independent hypothesis tests and were therefore interpreted according to the consistency of effect estimates across methods rather than subjected to additional multiple-testing correction. Concordance between MR-derived biomarker effects and pharmacodynamic responses to ruvonoflast was treated as a validation analysis and was not included within the multiple-testing correction framework. The positive- and negative-control analyses represented separate prespecified hypotheses, were not included in any FDR family, and were evaluated at a nominal (p<0.05) significance threshold.

### PheWAS Follow-up

A phenome-wide follow-up analysis was undertaken to systematically evaluate the clinical consequences of genetically proxied NLRP3 downregulation across binary outcomes from FinnGen Release 12 (500,348 individuals of Finnish ancestry)^79^ and its public meta-analyses with UK Biobank^80^ and with the Million Veteran Program and UK Biobank^71^.

Outcomes with fewer than 1,000 cases were excluded. Where an outcome was available from more than one of these sources, the source with the largest sample size was used, giving 1,387 binary outcomes (624 from FinnGen, 440 from the FinnGen–UK Biobank meta-analysis, and 326 from the FinnGen–Million Veteran Program–UK Biobank meta-analysis). Associations were estimated using IVW MR, using a fixed-effects model where there was no evidence of heterogeneity between variant-specific estimates (Cochran’s Q P≥0.05) and a multiplicative random-effects model where there was (Cochran’s Q P<0.05), or the Wald ratio where a single variant was available. Multiple testing was controlled using the false discovery rate (FDR) across all 1,387 outcomes (FDR-adjusted P<0.05). FDR-significant associations were further evaluated using weighted median and MR-Egger sensitivity analyses. Associations were considered robust when they demonstrated: (1) concordant direction of effect and P<0.05 using the weighted median estimator; (2) no evidence of directional horizontal pleiotropy by the MR-Egger intercept (P>0.05); and (3) where heterogeneity was present (Cochran Q P<0.05), concordant effect directions across IVW, weighted median, and MR-Egger estimates, with the MR-Egger estimate differing by no more than 20% from the IVW estimate.

### Software and reporting

Analyses were performed in R using the TwoSampleMR^81^, ieugwasr^82^, MendelianRandomization^83^, coloc^77^, MRPRESSO^75^, metafor^84^, and MungeSumstats^85^ packages. Reporting followed the STROBE-MR guidelines^86^.

## Ethics

The UK Biobank has obtained approval from the Northwest Multi-Center Research Ethics Committee (REC reference for UK Biobank is 11/NW/0382). All participants have provided written informed consent. The GWAS and eQTL summary statistics used in our study are provided by studies that received ethical approval and participant consent for both the analysis and distribution of summary-level data, as described in the original publications.

## Data Availability

All GWAS summary statistics are publicly available through their original repositories. Analytical code and processed outputs will be made publicly available upon publication.

## Results

### Instrument construction and biological validation

The primary instrument for genetically proxied NLRP3 downregulation comprised 14 CRP-lowering variants within the *NLRP3* locus (chromosome 1q44; **Supplementary Table S2**). Restricting the cis-window from ±100 kb to ±10 kb yielded an 11-variant instrument, which was used for sensitivity analyses **(Supplementary Table S2)**. A stricter r² threshold of <

0.01 yielded 5 variants in both ±100kb and ±10kb with 2 variants overlapping **(Supplementary Table S2)**. Across all cis-window and LD-clumping specifications, individual variant F statistics ranged from 34 to 408 **(Supplementary Table S2)**, indicating a low likelihood of weak-instrument bias. The 14- and 11-variant instruments explained 0.23% and 0.20% of the variance in serum CRP, respectively **(Supplementary Table S2)**. In the UK Biobank, median circulating CRP was 19% (95% CI, 14%-23%) lower among individuals in the lowest versus highest percentile of the weighted 14-variant genetic score **(Figure 2A; Supplementary Table S3)**. A concordant 19% reduction (95% CI, 14%-23%) was observed between the corresponding extremes of the restricted 11-variant score distribution **(Supplementary Table S3)**.

**Figure 2.**
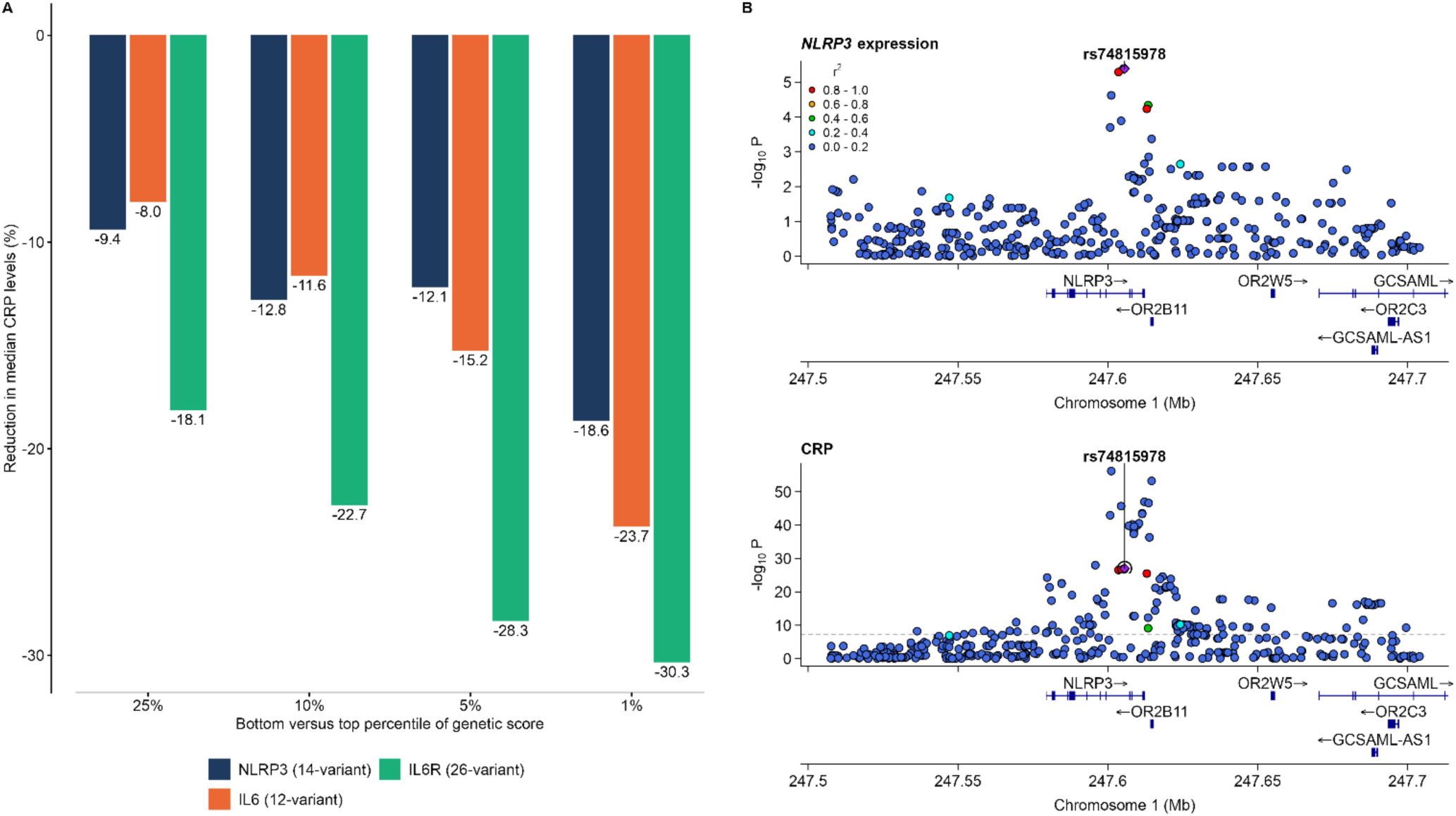
The NLRP3 genetic instrument is associated with measurable reductions in serum CRP and shares a causal variant with *NLRP3* expression in monocytes. **(A)** The effects of the 14-variant NLRP3 genetic score versus the previously described 12-variant IL6 and 26-variant IL6R genetic scores on reductions in CRP levels across percentiles. The bars represent the percentage reduction in median CRP levels between individuals in the bottom versus top 25%, 10%, 5% and 1% of each genetic score distribution in the UK Biobank (N = 464,264). **(B)** Regional association plots for *NLRP3* expression in naive monocytes (top; Fairfax et al., N = 414) and circulating CRP (bottom; N = 575,531) within a ±100-kb window of the *NLRP3* locus. Colocalization indicated rs74815978 as a shared causal variant (PP.H4 = 98.7%). The horizontal dashed line marks genome-wide significance (P = 5 × 10^−8^). IL-6: interleukin-6, IL-6R: interleukin-6 receptor, NLRP3: NOD-, LRR- and pyrin domain-containing protein 3, CRP: C-reactive protein.

None of the 14 instrument variants were located within the coding sequence of *NLRP3* **(Supplementary Table S4)**. However, the variants demonstrated cis-regulatory associations with *NLRP3* expression across whole blood, immune-cell populations, and nonimmune tissues **(Supplementary Figure 1; Supplementary Table S5)**. Ten of the 14 variants were genome-wide significant cis-eQTLs for *NLRP3* expression in whole blood. In the eQTL Catalogue, genome-wide significant cell-specific associations were predominantly observed in the myeloid lineage **(Supplementary Table S5)**. Consistent with the predominant activity of the NLRP3 inflammasome in monocytes and macrophages^42,43^, colocalisation in naïve monocytes demonstrated strong evidence that *NLRP3* expression and circulating CRP shared a causal variant (PP.H4=98.7%; **Figure 2B, Supplementary Table S6**).

### Downstream inflammatory pathway validation

In line with the known biological effects of NLRP3 inflammasome^43^, genetically proxied NLRP3 downregulation was associated with lower circulating IL-18 (β=−0.11; 95% CI, −0.14 to −0.08; P<0.0001) and IL-6 concentrations (β=−0.15; 95%CI, −0.17 to −0.12; P<0.0001) **(Figure 3A; Supplementary Table S7**). Both associations remained significant after correction for multiple testing and were directionally consistent across independent proteomic platforms **(Supplementary Table S7)**, weighted-median analyses, and analyses using the restricted 11-variant instrument **(Supplementary Tables S7 and S8)**, with no evidence of directional horizontal pleiotropy **(Supplementary Table S7)**. Estimates were robust to leave-one-out analysis **(Supplementary Table SG)**. Colocalisation provided strong evidence that the *NLRP3*-region association shared a causal variant with both IL-18 (PP.H4=99.1%) and IL-6 (PP.H4=99.6%; **Figure 3B; Supplementary Table S6**), further supporting that the instrument recapitulates the known role of NLRP3 in regulating downstream cytokine signaling, including IL-18 signaling and IL-1β-induced IL-6 production.

**Figure 3.**
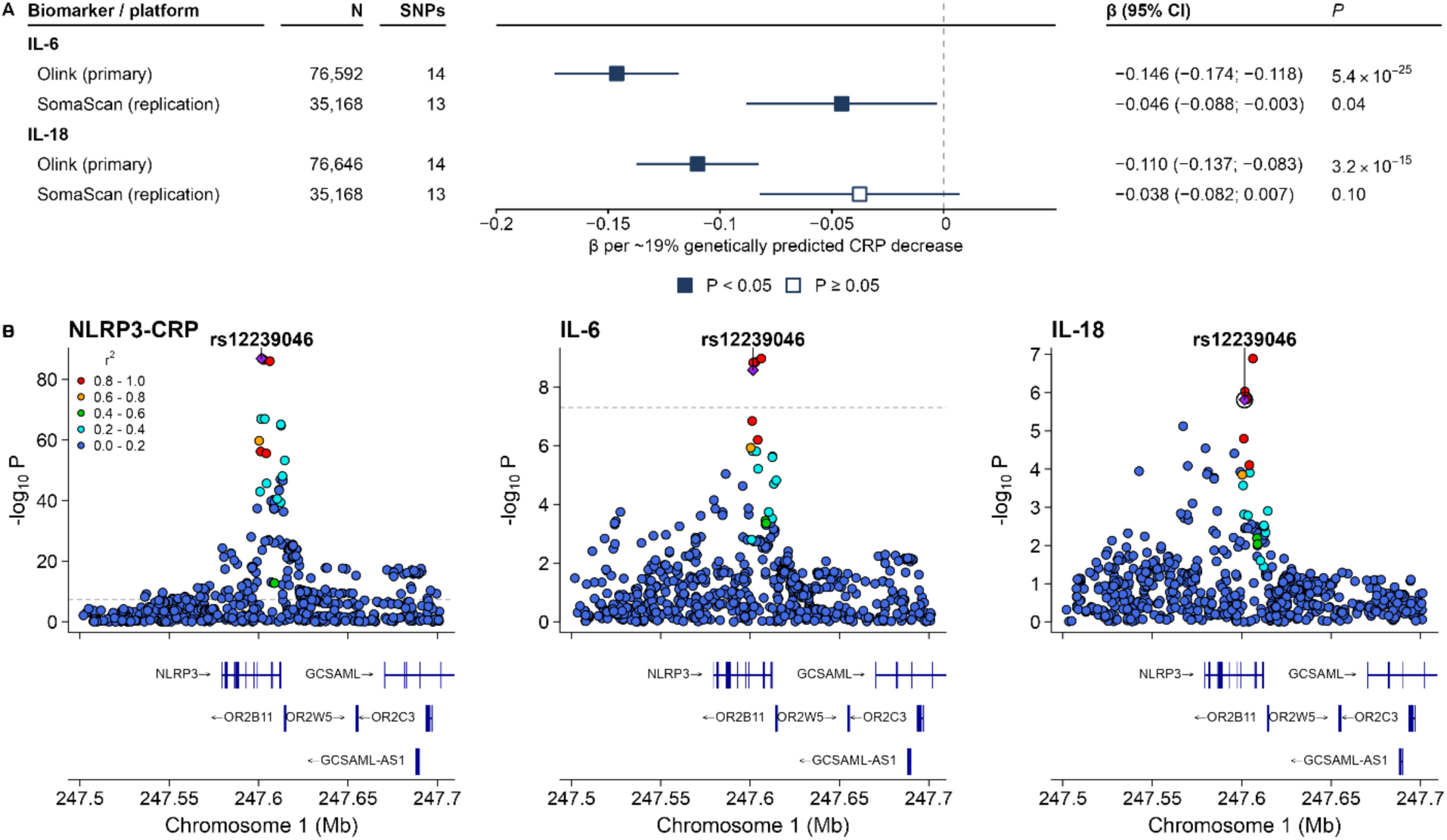
Genetically proxied NLRP3 downregulation is associated with lower circulating IL-6 and IL-18 levels. **(A)** IVW MR associations (fixed effects) of genetically proxied NLRP3 downregulation (14-variant instrument, scaled to a 19% decrease in CRP levels) with circulating IL-6 and IL-18, measured on the Olink platform (primary analysis; Koprulu et al.; IL-6 N = 76,592, IL-18 N = 76,646) and the SomaScan platform (deCODE; Ferkingstad et al.; N = 35,559 for each). **(B)** Regional association plots for CRP (NLRP3-CRP, left), IL-6 (middle) and IL-18 (right; Olink, Koprulu et al.) within a ±100-kb window of the *NLRP3* locus. Colocalization indicated rs12239046 as a shared causal variant for IL-6 (PP.H4 = 99.6%) and IL-18 (PP.H4 = 99.1%). The horizontal dashed line marks genome-wide significance (P = 5 × 10^−8^). All reported P values are two sided. IVW: inverse-variance weighted, SNP: single-nucleotide polymorphism, IL-6: interleukin-6, IL-18: interleukin-18, NLRP3: NOD-, LRR- and pyrin domain-containing protein 3, CRP: C-reactive protein, CI: confidence intervals.

Furthermore, the biomarker effects of genetically proxied NLRP3 downregulation were broadly consistent in direction with the pharmacodynamic response to ruvonoflast, a small molecule NLRP3 inhibitor, as tested in a randomized, double-blind phase 1b trial^13^, with similar findings across the 14- and 11-variant instruments **(Figure 4A and 4B; Supplementary Tables S8 and S10)**. Except for IL-18 that was reduced by our instrument but paradoxically not by ruvonoflast, there were broadly consistent effects across 7 biomarkers, including IL-6, lipoprotein(a), fibrinogen, triglycerides, total cholesterol, LDL cholesterol, and HDL cholesterol **(Figure 4C)**. Although, rank correlation was non-significant (ρ = 0.64; P = 0.094). Together, these results supported the validity of the genetic instrument as a proxy of NLRP3 inhibition.

**Figure 4.**
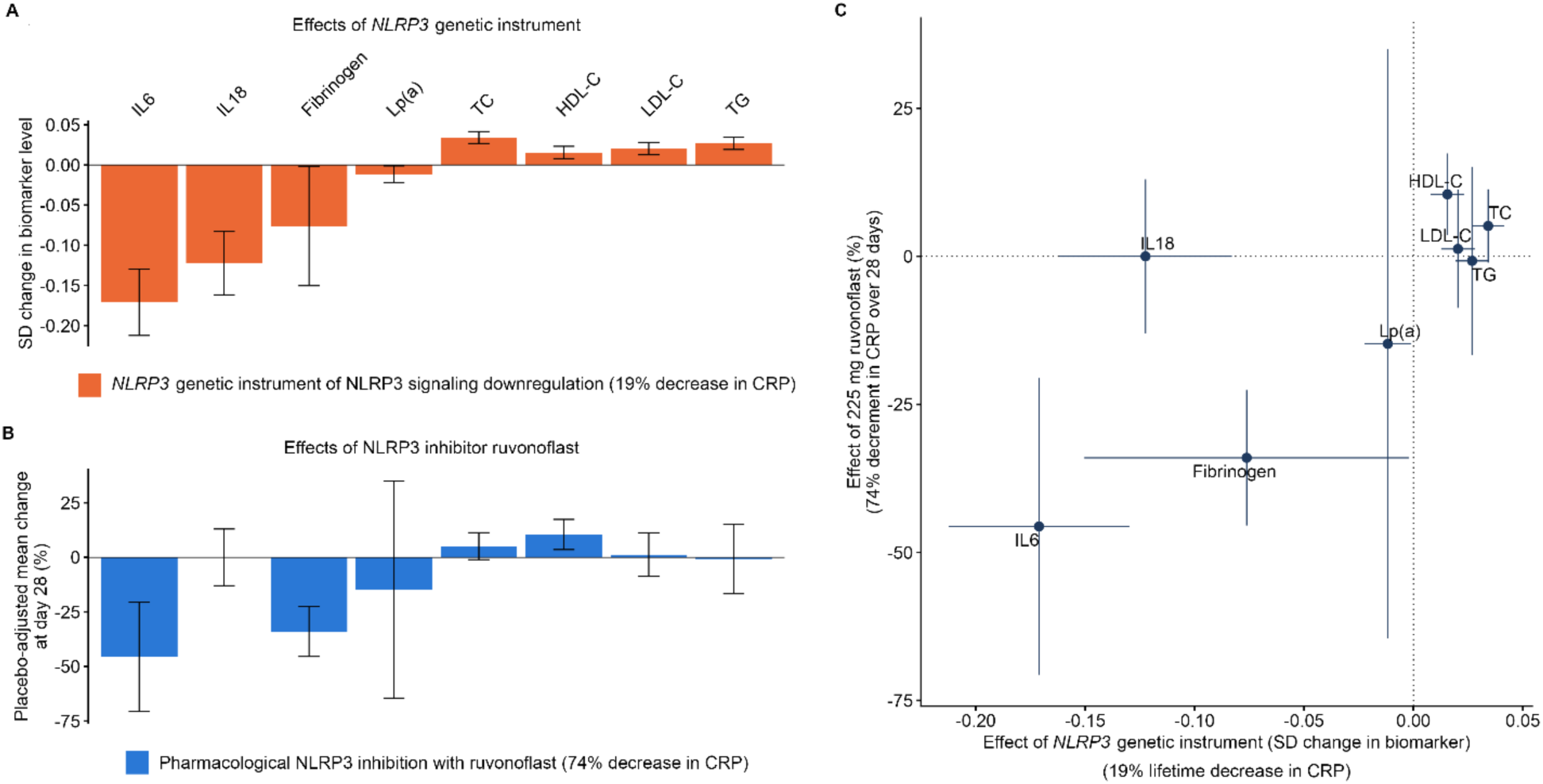
Concordance between the biomarker effects of genetically proxied NLRP3 downregulation and pharmacological NLRP3 inhibition. **(A)** IVW MR associations of genetically proxied NLRP3 downregulation (14-variant instrument, scaled to a 19% decrease in CRP levels) with 8 circulating biomarkers measured in a randomized, double-blind phase 1b trial of ruvonoflast (oral NLRP3 inhibitor). Data are presented in SD units of each biomarker and error bars correspond to 95% CIs. **(B)** The effects of ruvonoflast 225 mg twice daily versus placebo on the 28-day change in the same 8 biomarkers (which led to a 74% decrease in high-sensitivity CRP over 28 days). Data are presented as the placebo-adjusted mean percentage change from baseline with 95% CI. **(C)** Correlation of the effects of the NLRP3 genetic instrument (scaled to a 19% decrease in CRP; n = 575,531) and ruvonoflast (225 mg versus placebo) on the 8 biomarkers. Each point represents an effect estimate for a biomarker, as derived by IVW MR, with horizontal error bars indicating the 95% CIs for the genetic instrument effects. CRP: C-reactive protein, HDL-C: high-density lipoprotein cholesterol, IL-6: interleukin-6. IL-18: interleukin-18, LDL-C: low-density lipoprotein cholesterol, Lp(a): lipoprotein(a), NLRP3: NOD-, LRR- and pyrin domain-containing protein 3, TC: total cholesterol, TG: triglycerides.

### Cardiovascular and cardiometabolic outcomes

Next, we evaluated associations of genetically proxied NLRP3 downregulation with clinical cardiovascular and cardiometabolic outcomes relevant to therapeutic development efforts for NLRP3 inhibitors. Consistent with the established efficacy of pharmacological IL-1β inhibition in pericarditis^55,87,88^, genetically proxied NLRP3 downregulation was associated with a lower risk of recurrent pericarditis (OR per 19% reduction in CRP, 0.68; 95% CI, 0.58–0.80; **Figure 5A; Supplementary Table S11)**. However, among the primary clinical outcomes, there was no evidence of reduced risk of coronary artery disease (OR, 1.06; 95% CI, 1.03–1.08), peripheral arterial disease (OR, 1.10; 95% CI, 1.04–1.15), or type 2 diabetes (OR, 1.03; 95% CI, 1.01–1.04) with effects pointing to the opposite direction **(Figure 5A; Supplementary Table S11)**. There was also no association with heart failure (OR, 1.00; 95% CI, 0.97–1.02) or ischaemic stroke (OR, 0.98; 95% CI, 0.94–1.02). Effect estimates were directionally consistent in weighted-median analyses, and leave-one-out analyses **(Supplementary Tables S11 and S12),** whereas MR-Egger estimates were attenuated and less precise, with no evidence of directional horizontal pleiotropy (MR-Egger intercept P>0.05 for each outcome; **Supplementary Tables S11)**. For outcomes that were significant in the primary analysis, findings were similar using the restricted 11-variant instrument **(Supplementary Table S8)**. Colocalisation provided little evidence that the *NLRP3*-region signal shared a causal variant with recurrent pericarditis (PP.H4=2%) or the primary cardiometabolic outcomes **(Supplementary Table S6)**. There was no association with the negative-control outcome of left-handedness (P=0.54; **Supplementary Table S11**).

**Figure 5.**
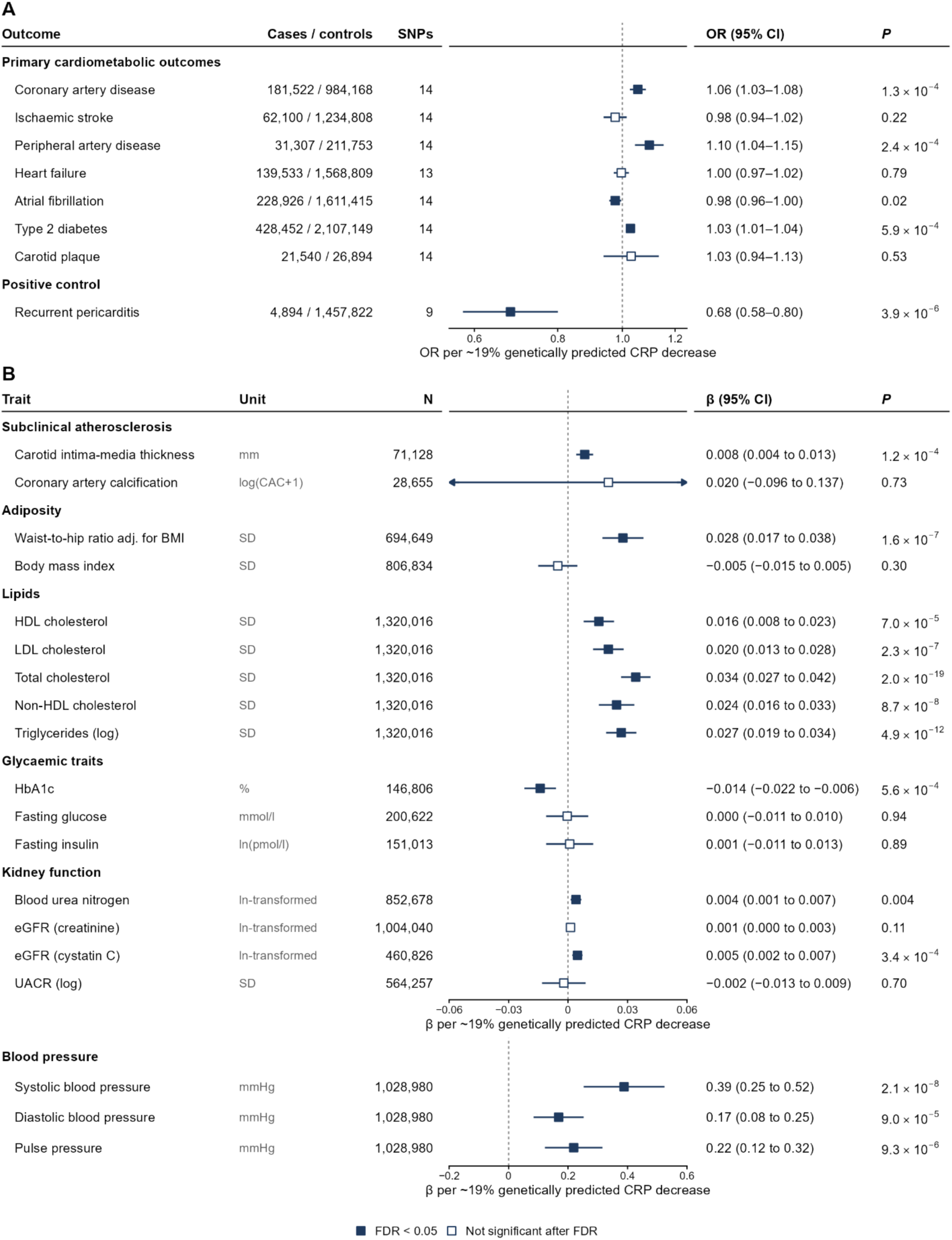
Genetically proxied NLRP3 downregulation is associated with lower risk of recurrent pericarditis but not with lower risk of atherosclerotic cardiovascular disease. IVW MR associations (fixed effects) of genetically proxied NLRP3 downregulation (14-variant instrument, scaled to a 19% decrease in CRP levels) with **(A)** clinical cardiovascular and cardiometabolic endpoints and with recurrent pericarditis (positive control) and **(B)** subclinical atherosclerosis, blood pressure, adiposity, lipid, glycemic and renal traits. Results are presented as effect estimates per 19% decrease in CRP, in the units of each source GWAS, and error bars correspond to 95% CIs. All reported P values are two sided. SNP: single nucleotide polymorphism, OR: odds ratio, CI: confidence intervals, CRP: C-reactive protein, CAC: coronary artery calcification, BMI: body mass index, SD: standard deviation, ln: natural logarithm, HDL: high density lipoprotein, LDL: low density lipoprotein, HbA1c: glycated hemoglobin, eGFR: estimated glomerular filtration rate, UACR: urinary albumin-to-creatinine ratio, FDR: false discovery rate

Among secondary exploratory outcomes, genetically proxied NLRP3 downregulation was associated with higher carotid intima-media thickness (β=0.01; 95% CI, 0.004–0.013; **Figure 5B; Supplementary Table S11**), higher waist-to-hip ratio adjusted for BMI (β=0.13; 95% CI, 0.017–0.038), and higher levels of systolic and diastolic blood pressure, pulse pressure, HDL, LDL, total and non-HDL cholesterol, and triglycerides (**Supplementary Table S11**). Colocalisation provided evidence of a shared causal variant between the NLRP3-region signal for CRP and total cholesterol (PP.H4=92.4%). Evidence for colocalization with systolic blood pressure did not meet the threshold (PP.H4=49.6%), while no evidence of colocalisation was observed for diastolic blood pressure, carotid intima-media thickness, or HDL cholesterol (**Supplementary Table S6**).

### Infectious disease outcomes

Given the increased incidence of serious infections observed with IL-1β inhibition in CANTOS^5–7^, we evaluated associations of the constructed instrument with infectious disease outcomes. Genetically proxied NLRP3 downregulation was associated with lower odds of pneumonia requiring hospital admission (OR, 0.93; 95% CI, 0.90–0.96; P=8.5×10⁻⁵), the only prespecified infectious outcome that remained significant after FDR correction **(Supplementary Figure 2; Supplementary Table S13)**. There was no evidence of heterogeneity (Cochran Q P=0.88) or directional horizontal pleiotropy (MR-Egger intercept P=0.75; **Supplementary Table S13**), and the direction of association was consistent in weighted-median (OR, 0.95; P=0.028), MR-PRESSO analyses (OR, 0.93; P<0.001), leave-one-out analyses and the 11-variant instrument **(Supplementary Tables S8, S13 and S14)**. However, colocalisation provided little evidence that the NLRP3-region and pneumonia association signals shared a causal variant (PP.H4=0.6%; **Supplementary Table S6)**. We observed no associations with higher risk of any other of the tested infectious disease outcomes.

### Phenome-Wide Analysis

In a phenome-wide associations study, we explored associations between genetically proxied NLRP3 inhibition and 1,387 binary clinical outcomes with more than 1,000 cases. After correcting for multiple comparisons (false discovery rate (FDR)-corrected P < 0.05) and excluding outcomes that lacked consistency across sensitivity MR methods, we found 94 outcomes to be significantly associated with genetically proxied NLRP3 downregulation **(Supplementary Table S15; Figure 6).** Genetically proxied NLRP3 downregulation was associated with higher odds of the following clinical outcomes: (1) aortic aneurysm (abdominal aortic aneurysm and aortic aneurysm, broader definition), (2) peripheral vascular disease, (3) dyslipidemia (for example, statin use, hypercholesterolemia and disorders of lipoprotein metabolism), (4) diabetes mellitus, (5) non-alcoholic fatty liver disease, (6) hypertrophic cardiomyopathy, (7) dementia, (8) glaucoma and (9) musculoskeletal disease (for example, rotator cuff syndrome, Dupuytren’s contracture and lateral epicondylitis). However, genetically proxied NLRP3 inhibition was associated with lower odds of: (1) tonsillar disease and upper and lower respiratory tract infections (for example, chronic diseases of tonsils and adenoids, tonsillectomy, acute upper respiratory infections and pneumonia), (2) appendicitis, (3) macular degeneration, (4) heart failure, (5) non-small cell lung cancer, (6) intrahepatic cholestasis of pregnancy, (7) reactive arthropathies, (8) erythema nodosum and lichen simplex, and (9) asthma **(Figure 6 and Supplementary Table S15).** These signals highlight potential repurposing opportunities or safety concerns that might warrant further investigation.

**Figure 6.**
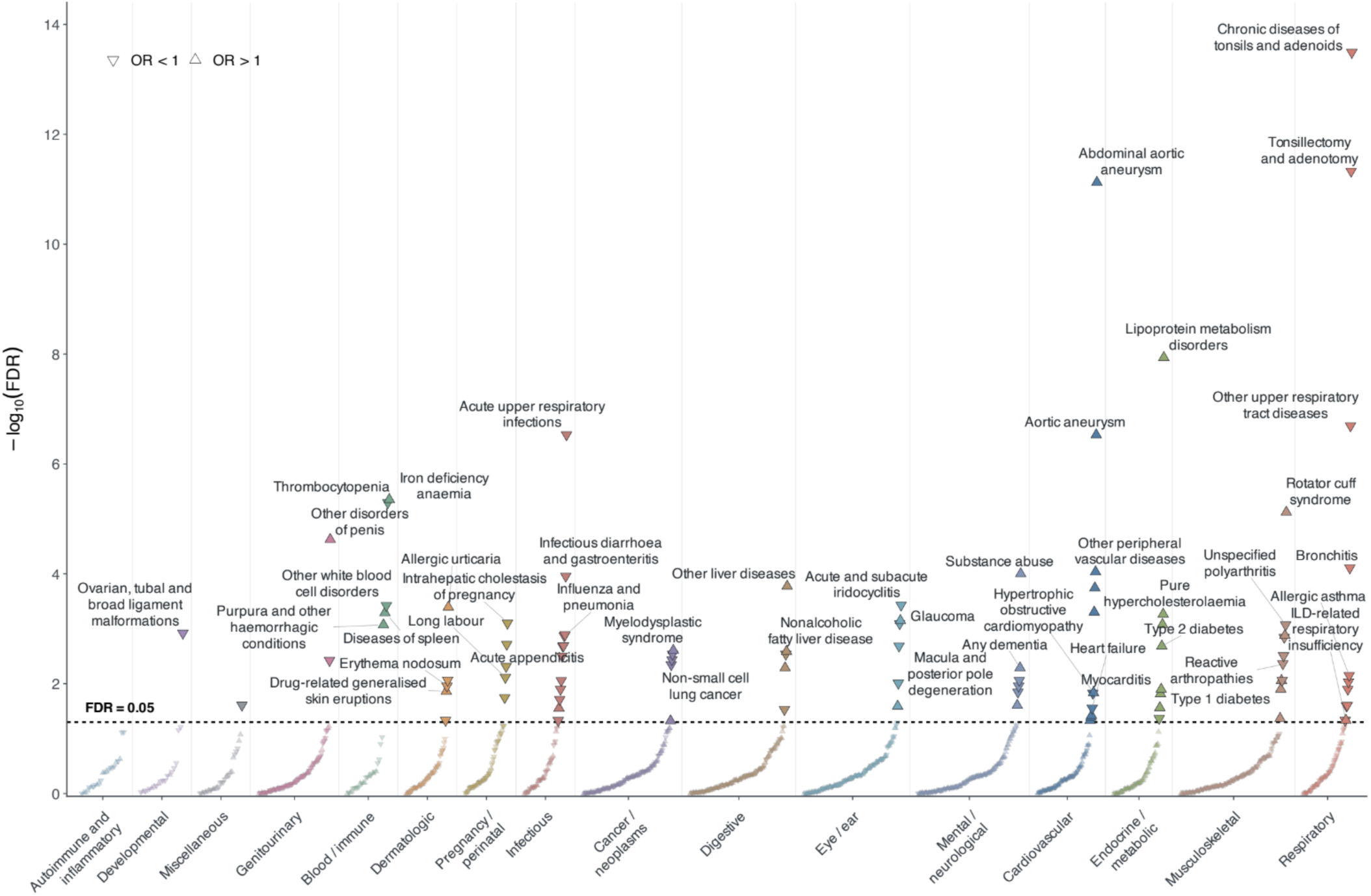
PheWAS of genetically proxied NLRP3 downregulation highlights repurposing opportunities and safety signals. Phenome-wide association study (PheWAS) evaluating the effects of genetically proxied NLRP3 downregulation across 1,387 binary clinical outcomes with more than 1,000 cases in the population-based FinnGen cohort. Associations were estimated using inverse-variance weighted Mendelian randomization (IVW MR). Downward and upward arrows indicate associations with lower and higher odds of clinical outcomes, respectively. Annotated outcomes met the prespecified significance and robustness criteria based on sensitivity analyses, as detailed in the Methods. The horizontal dashed line indicates the FDR threshold of 0.05. The y-axis represents the negative base-10 logarithm of FDR-adjusted P values. ILD: interstitial lung disease, OR: odds ratio, FDR: false discovery rate

## Discussion

In this study, we developed a cis-genetic instrument for NLRP3 downregulation that reproduced the expected molecular consequences of NLRP3 inhibition but did not support a protective effect on ASCVD. Instrument validity was supported by effects on *NLRP3* expression, strong colocalisation between *NLRP3* expression and circulating CRP, reductions in downstream IL-18 and IL-6, and concordance with the pharmacodynamic profile of the NLRP3 inhibitor ruvonoflast. Genetically proxied NLRP3 downregulation was also associated with a lower risk of recurrent pericarditis, consistent with the established therapeutic effect of IL-1β pathway inhibition. Despite this biological validation, NLRP3 downregulation was not associated with protection against any major ASCVD phenotypes, with modestly higher odds of CAD and PAD alongside several adverse cardiometabolic traits. We found no associations with increased risk of any infectious disease endpoints.

Finally, phenome-wide analyses largely reinforced the absence of a protective cardiovascular signal and identified additional associations of potential relevance to safety and therapeutic repurposing.

These findings have implications for the evolving therapeutic landscape of cardiovascular inflammation. CANTOS established proof of concept that targeting inflammation through IL-1β inhibition can reduce cardiovascular events^5–7^, while subsequent genetic and pharmacological evidence has supported the downstream IL-6 pathway as a therapeutic target^8–10,17^. However, the absence of cardiovascular efficacy in ZEUS and discontinuation of the HERMES and ATHENA programmes have introduced uncertainty regarding the extent to which suppression of this pathway translates into clinical cardiovascular benefit^11,89,90^.

NLRP3 represents a more proximal target, regulating inflammasome-dependent maturation of IL-1β and IL-18, and several small-molecule inhibitors are in clinical development^91^. Ruvonoflast reduced hsCRP and downstream inflammatory biomarkers, but whether these pharmacodynamic effects translate into fewer cardiovascular events remains unknown^13^. Our observation that genetically proxied NLRP3 downregulation reproduced key features of this pharmacodynamic profile without reducing ASCVD risk suggests that biological target engagement and suppression of circulating inflammatory biomarkers alone may be insufficient to predict cardiovascular efficacy.

The divergence between the molecular and cardiovascular effects of NLRP3 downregulation may partly reflect its position within the inflammatory cascade. Unlike IL-6, which directly stimulates hepatic CRP production, NLRP3 lies further upstream, with its effects propagated through inflammasome-dependent maturation of IL-1β and subsequent IL-6 signalling^92^. Consistent with this greater biological distance, the *NLRP3* genetic score produced an approximately 19% difference in circulating CRP, whereas the *ILC* and *ILCR* instruments developed by Zhang et al. produced 24% and 30% differences, respectively, and were associated with lower risks of CAD, PAD, and large artery atherosclerotic stroke^17^. These estimates are not directly comparable across instruments, but they illustrate that similar reductions in a distal inflammatory biomarker need not imply equivalent perturbation of the underlying pathway or equivalent cardiovascular effects.

Genetically proxied NLRP3 inhibition was associated with small increases in circulating lipid markers, blood pressure and waist-hip ratio measurements, as well as slightly elevated risk of type 2 diabetes. This raises the possibility that an adverse cardiometabolic profile may offset benefits from immunomodulation. However, these associations were not reproduced during 28 days of pharmacological NLRP3 inhibition with ruvonoflast. Despite hsCRP suppression, reductions in body weight, total cholesterol, LDL cholesterol and triglycerides were comparable between the intervention and placebo groups. Similarly, a Phase 1b study of dapansutrile, oral NLRP3 inhibitor, in patients with heart failure with reduced ejection fraction (n = 24) identified small reductions in blood pressure measurements after approximately 12 days^93^. Both studies are limited by their short treatment durations, small samples and in the ruvonoflast trial, concurrent calorie restriction. The divergence between genetic and pharmacological findings may reflect metabolic consequences of lifelong *NLRP3* downregulation that are not reproduced by inhibition initiated in adulthood. Such effects could potentially contribute to the absence of genetically predicted ASCVD protection without necessarily excluding cardiovascular benefit from pharmacological NLRP3 inhibition. Moreover, our instrument was associated with increased risk of type 2 diabetes, despite being associated with minimal, albeit significant, decreases in HbA1c levels in a population without diabetes. This discrepancy may reflect genetic effects on type 2 diabetes risk through pathways not adequately captured by fasting glycaemic parameters or may be attributable to horizontal pleiotropy.

Lifelong genetic perturbation may also differ importantly from pharmacological NLRP3 inhibition. Chronic downregulation could induce compensatory responses within the innate immune network that are not reproduced by shorter-term therapeutic inhibition. Experimental evidence in mice that *Nlrp3* haploinsufficiency is accompanied by upregulation of *Nlrp1* provides one potential example of such pathway compensation^31^. Although these mechanisms cannot be established from the present genetic analyses, they provide a plausible explanation for why lifelong genetically proxied NLRP3 downregulation could reproduce expected downstream biomarker effects without conferring the anticipated cardiovascular benefit. Conversely, this distinction means that our findings should not be interpreted as excluding cardiovascular benefit from pharmacological NLRP3 inhibition. The recent Lp(a)HORIZON trial provides a parallel example of potential discordance between genetic and pharmacological evidence, where substantial pharmacological Lp(a) lowering failed to reduce cardiovascular events despite robust genetic evidence supporting a causal role for Lp(a) in ASCVD^94^, highlighting the limitations of extrapolating lifelong genetic effects to therapeutic interventions initiated later in life.

We found no evidence that genetically proxied NLRP3 downregulation increased susceptibility to prespecified infectious outcomes. This contrasts with the increased risk of serious or fatal infection observed with downstream IL-1β inhibition in CANTOS^5–7^ and raises the possibility that targeting NLRP3 may have distinct effects on host defence compared with broader suppression of IL-1β signalling. Notably, genetic NLRP3 downregulation was associated with lower odds of pneumonia requiring hospital admission, with directionally consistent estimates across sensitivity analyses. However, the limited colocalisation support suggests that this association may not be attributable to the same causal variant underlying genetically proxied NLRP3 downregulation and therefore warrants cautious interpretation. More broadly, genetic analyses of acute infection are inherently influenced by pathogen exposure, healthcare-seeking behaviour, and case ascertainment and may not capture the consequences of short-term pharmacological inhibition during an infectious challenge. These findings therefore provide some reassurance against a large increase in infection susceptibility but should not be interpreted as establishing the infectious safety of pharmacological NLRP3 inhibition, which will require prospective evaluation in adequately powered clinical trials.

Finally, phenome-wide analyses identified potential safety signals and therapeutic opportunities beyond the prespecified outcomes. Associations with aortic aneurysm, peripheral vascular disease, diabetes, non-alcoholic fatty liver disease, dementia, glaucoma, and hypertrophic cardiomyopathy warrant targeted investigation during NLRP3 inhibitor development, while inverse associations with heart failure and non-small-cell lung cancer may suggest opportunities for therapeutic repurposing. These findings are hypothesis-generating and require independent replication and mechanistic or pharmacological validation. The inverse association with heart failure is noteworthy, particularly given the absence of evidence for cardiovascular benefit across the major atherosclerotic outcomes. Although the primary heart failure analysis was null, the larger PheWAS analysis yielded an effect estimate in the same protective direction, suggesting that the discrepancy may partly reflect differences in statistical power. This signal is supported by limited early clinical evidence. A pooled analysis of dapansutrile in patients with heart failure with reduced ejection fraction (n = 24) demonstrated an approximately 2% improvement in left ventricular ejection fraction after 12 days^93^. However, treatment was not associated with reductions in hsCRP or NT-proBNP, and the small sample size, short treatment duration, and early-phase design preclude conclusions regarding clinical efficacy^93^. Collectively, the genetic and preliminary pharmacological findings raise the possibility that the effects of NLRP3 inhibition may differ between heart failure and atherosclerotic cardiovascular disease, but whether pharmacological NLRP3 inhibition improves cardiac function or clinical outcomes in HFrEF remains uncertain.

Several limitations should also be considered. Mendelian randomization estimates the consequences of relatively small, lifelong genetically determined differences in pathway activity and cannot directly reproduce the magnitude, timing, reversibility, tissue specificity, or treatment context of pharmacological NLRP3 inhibition^32^. Consequently, the absence of a genetically predicted cardiovascular benefit does not establish that pharmacological NLRP3 inhibition cannot reduce cardiovascular events. Although the instrument showed strong biological and pharmacodynamic validation, CRP is a distal readout of NLRP3 activity, and the selected variants were noncoding, potentially influencing the expression of neighbouring genes. Horizontal pleiotropy therefore cannot be excluded; however, nearby genes have no established roles in inflammasome signaling and there was significant colocalization between effects on CRP and *NLRP3* expression in monocytes, reducing the biological plausibility of alternative mechanisms^95^. The limited colocalisation observed for several clinical outcomes further warrants caution when attributing these associations specifically to NLRP3 perturbation. Most contributing GWASs were conducted predominantly in individuals of European ancestry, limiting generalisability to other ancestral populations. Finally, phenome-wide analyses are inherently exploratory and, despite multiple-testing correction and sensitivity analyses, require independent replication before individual associations can be considered established therapeutic or safety effects.

In conclusion, we developed a genetic instrument for NLRP3 downregulation that recapitulated the established biology of the NLRP3 inflammasome and key molecular effects of pharmacological NLRP3 inhibition. Still, genetically proxied NLRP3 downregulation was not associated with reduced ASCVD risk or a more favorable cardiometabolic risk profile.

## Supporting information

Supplementary tables, figures and MR-STROBE checklist

## Data Availability

All GWAS summary statistics are publicly available through their original repositories.

## Acknowledgments

Data for the UK Biobank analyses were accessed under application number 151281.

## Supplemental Material

Tables ST1-ST15

Figures S1-S2

STROBE-MR reporting guideline

