## Supplementary tables, figures and MR-STROBE checklist for "Genetically Proxied NLRP3 Inflammasome Downregulation and Cardiovascular Disease": nlrp3_supp_figures_medrxiv.pdf

### Supplementary figures

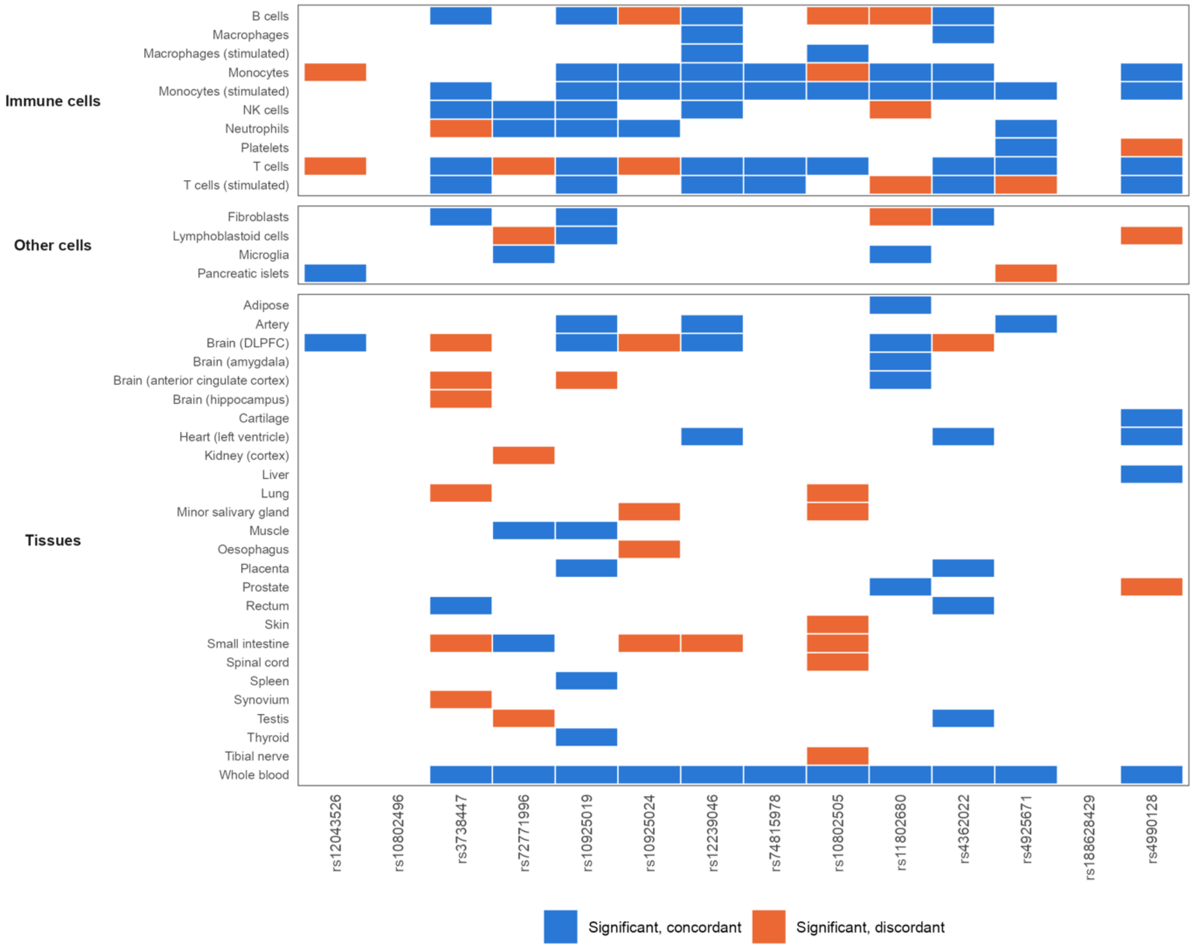


**Supplementary Figure S1. *NLRP3* cis-eQTL associations of the NLRP3 instrument variants across all eQTL datasets and conditions.** The direction and significance of the association of each NLRP3 instrument variant (columns, ordered by genomic position) with *NLRP3* expression across every dataset in the eQTL Catalogue and eQTLGen. Directions are expressed relative to the CRP-lowering allele. Blue boxes indicate nominally significant (two-sided P < 0.05) associations of the CRP-lowering allele with lower *NLRP3* expression (concordant); orange boxes indicate nominally significant associations with higher expression (discordant). eQTL: expression quantitative trait loci, DLPFC: dorsolateral prefrontal cortex

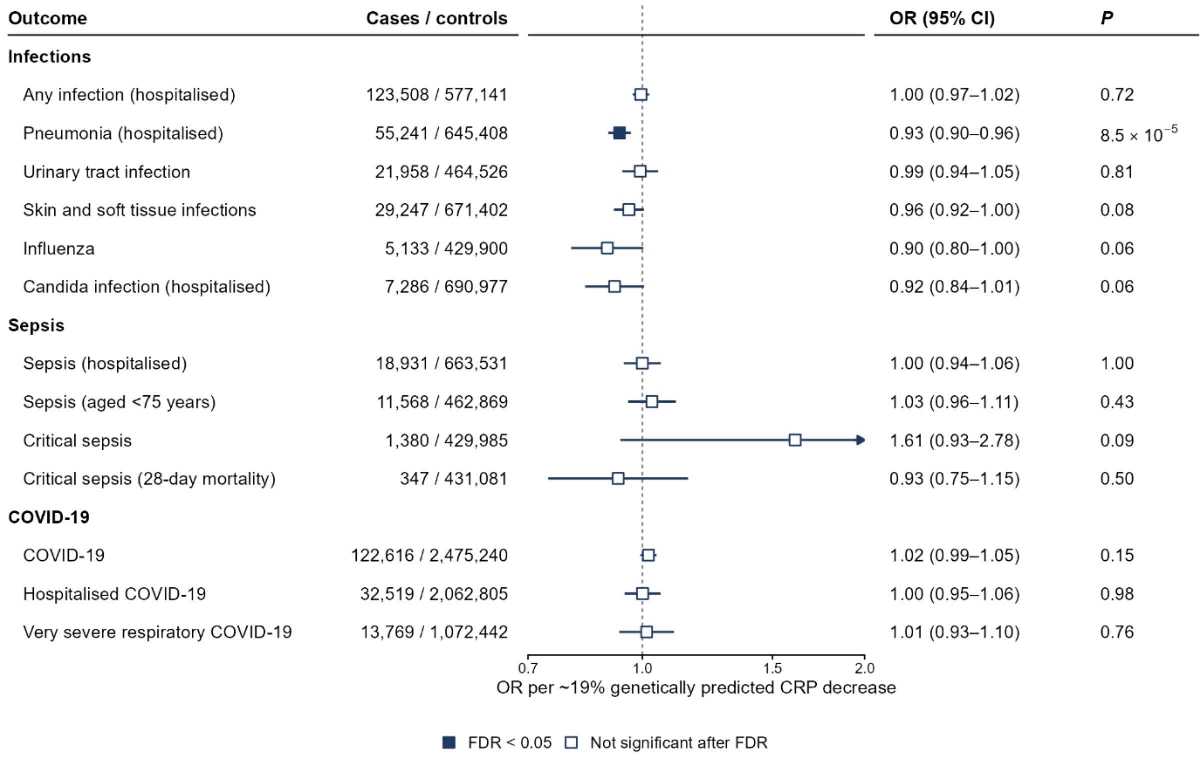


**Supplementary Figure S2. Effects of genetically proxied NLRP3 downregulation on infectious disease outcomes.** IVW MR associations (fixed effects) of genetically proxied NLRP3 downregulation (14-variant instrument, scaled to a 19% decrease in CRP levels) with clinical endpoints related to hospital-treated infections, sepsis and viral infections (COVID-19 and influenza). Data are presented as ORs per 19% decrease in CRP and error bars correspond to 95% CIs. CRP: C-reactive protein, OR: odds ratio, CI: confidence intervals, FDR: false discovery rate
